# Long-term survival analysis of HIV patients on antiretroviral therapy in Congo: a 14 years retrospective cohort analysis, 2003-2017

**DOI:** 10.1101/2022.03.15.22272252

**Authors:** Gilbert Ndziessi, Ange Clauvel Niama, Arkadit Jeandria Nkodia, Merlin Diafouka

## Abstract

**Background:** The long-term survival of patients on antiretroviral treatment in Congo remains less documented. Our study aimed to analyze the long-term survival of adults living with HIV on ART (Antiretroviral Therapy).

**Methods:** We conducted a historical cohort study on 2,309 adult PLHIV (People Living with HIV) followed between January 1, 2003 and December 31, 2017 whose viral load and date of initiation of ART were known. The Kaplan Meier method was used to estimate the probability of survival and the Cox regression model to identify factors associated with death.

**Results:** The median age was 49 years; the female sex was predominant with 68.56%. The probability of survival at 14 years was 83%, (95% CI (Confidence Interval) [78-87]). On the other hand, when the lost to follow-up died, it was 66% (95% CI [62-70]) in the worst scenario. Stratified cox regression analysis showed that: being male, AHR (Adjusted Hazard Ratio) = 1.65 (95% CI [1.26-2.17]) was significantly associated with death, p-value <0.0001. Furthermore, having a viral load> 1000 copies / ml, AHR = 2.56 (95% CI [1.93-3.40]), be in the advanced WHO clinical stage, in particular: stage II, AHR = 4.07 (95% CI [2.36-7.01]); stage III, AHR = 13.49 (95% CI [8.99-20.27]) and stage IV, AHR = 34.45 (95% CI [23.74-50]) were also significantly associated with death; p-value <0.0001.

**Conclusion:** The long-term survival of PLHIV is worrying despite the offer of ARVs.

## Introduction

HIV / AIDS remains a major public health concern in the world, particularly in low and middle-income countries [1]. Since the start of the epidemic, 77.3 million [59.9 million - 100 million] people have been infected with HIV. In 2017, 36.9 million [31.1 million - 43.9 million] people were living with HIV, including 1.8 million [1.4 million - 2.4 million] new infections [2]. The African Region of the World Health Organization (WHO), where 25.7 million people were living with HIV in 2017, is the most affected region. It also concentrates more than two-thirds of new infections occurring in the world with over 35 million deaths worldwide [3]. According to the WHO, between 2000 and 2016, 13.1 million lives were saved thanks to antiretroviral treatment (ART). Access to these treatments in sub-Saharan Africa remains very low due to many obstacles [4, 5], such as the low number of physicians, limited access to viral load measurement and the low availability of basic tests such as measurement of TCD4 cells or biochemistry tests [6]. Faced with these difficulties, WHO has proposed a “lean” approach to support large-scale access to ART in countries with limited resources. [7]. These programs have shown results comparable to those obtained in northern countries in terms of survival, compliance, emergence of resistance, toxicity, virological, immunological and clinical efficacy [8, 9].

In the Republic of Congo, the prevalence of HIV in the population aged 15 to 49 is estimated at 3.2% [10]. This country developed an early response to reduce the spread of the virus from the first years of the discovery of the first cases in 1983. The establishment of a committee to diagnose and fight against HIV AIDS in 1985 and a National AIDS Control Program (PNLS) in 1987 followed this. A National Blood Transfusion Center (CNTS) and an Ambulatory Treatment Center (CTA) were created in Brazzaville in 1994. Since 2002, this national response has been marked by a significant commitment, with the establishment of a National AIDS Control Council (CNLS) [11].

In Congo, The treatment regimens for the management of HIV patients in Congo are classified into two periods: Before (Guidelines published in 2014) and after (Guidelines published in 2018). The details are in Table 1.

**Table 1.**
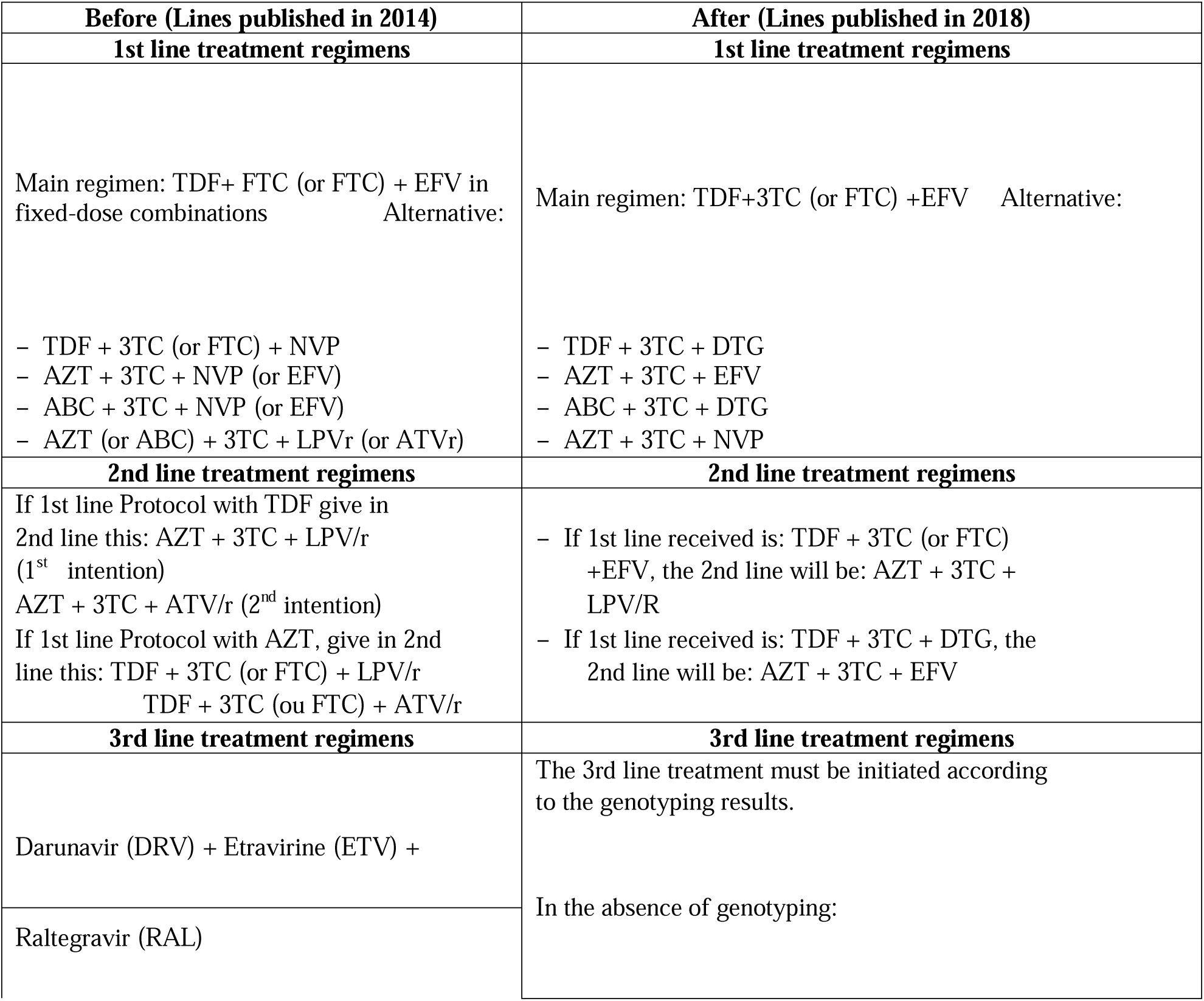
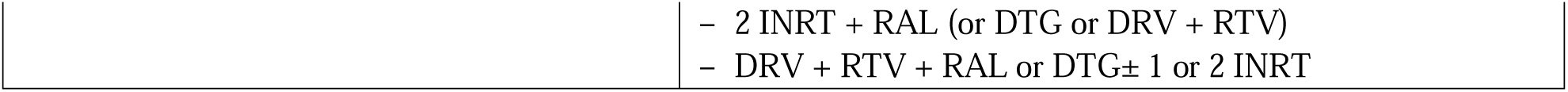
Overview of therapeutic regimens for providing care to HIV patients, Republic of Congo.

| **Before (Lines published in 2014)** | **After (Lines published in 2018)** |
| --- | --- |
| **1st line treatment regimens** | **1st line treatment regimens** |
| Main regimen: TDF+ FTC (or FTC) + EFV in fixed-dose combinations Alternative:<br>– TDF + 3TC (or FTC) + NVP<br>– AZT + 3TC + NVP (or EFV)<br>– ABC + 3TC + NVP (or EFV)<br>– AZT (or ABC) + 3TC + LPVr (or ATVr) | Main regimen: TDF+3TC (or FTC) +EFV Alternative:<br>– TDF + 3TC + DTG<br>– AZT + 3TC + EFV<br>– ABC + 3TC + DTG<br>– AZT + 3TC + NVP |
| **2nd line treatment regimens** | **2nd line treatment regimens** |
| If 1st line Protocol with TDF give in 2nd line this: AZT + 3TC + LPV/r (1<sup>st</sup> intention)<br>AZT + 3TC + ATV/r (2<sup>nd</sup> intention)<br>If 1st line Protocol with AZT, give in 2nd line this: TDF + 3TC (or FTC) + LPV/r<br>TDF + 3TC (ou FTC) + ATV/r | – If 1st line received is: TDF + 3TC (or FTC) +EFV, the 2nd line will be: AZT + 3TC + LPV/R<br>– If 1st line received is: TDF + 3TC + DTG, the 2nd line will be: AZT + 3TC + EFV |
| **3rd line treatment regimens** | **3rd line treatment regimens** |
| Darunavir (DRV) + Etravirine (ETV) +<br>Raltegravir (RAL) | The 3rd line treatment must be initiated according to the genotyping results.<br>In the absence of genotyping: |

|  |
| --- |
| - 2 INRT + RAL (or DTG or DRV + RTV)<br>- DRV + RTV + RAL or DTG± 1 or 2 INRT |

Available data shows that over 80% of patients start treatment late and viral load is often uncontrolled [12]. Loss of follow-up is common, and medication adherence is estimated between 55.4% and 86.9% [13]. Other results revealed compliance rates of 84% during the first 12 months as well as irregular use of ARVs. [14, 15]. These conditions can increase the mortality of PLHIV. In addition, few studies have been done to document these aspects. This study aims to fill these gaps by using the Eval-Co cohort database. It aimed to estimate the probability of survival at 14 years while identifying the prognostic factors associated with the death of PLHIV on ART between 2003 and 2017 in Congo.

## Methods

### Study type and period

We performed a historical cohort study, including patients on ART between 2003 and 2017, in the Republic of Congo.

### Presentation of the data source: Eval-CO cohort

The data used for our study come from the historical cohort 2003-2017 of the EVAL-CO project (evaluation of the therapeutic management of HIV patients in Congo).

The primary objective of EVAL-CO was to assess the survival of HIV patients after initiation of treatment in Congo. Secondary objectives were to: 1) assess retention of patients on antiretroviral therapy, 2) measure the proportions of patients with CD4 cell counts below <350 cells/mm3 among those with baseline CD4 cell counts, 3) assess the virological efficacy of treatment regimens, and 4) measure the incidence of tuberculosis and hepatitis B among people living with HIV in Congo. The Faculty of Health Sciences led this project. The Eval-Co database of the retrospective cohort was formed from fragmented data from the various centralized health facilities at the level of the CNLS-E (national council for the fight against AIDS and epidemics) and the CTA (Outpatient Treatment Center) of Brazzaville, Pointe-Noire and other departments of the Republic of Congo. It integrates the data of HIV positive patients at the initiation of treatment (Visit 0) and the data collected during the various visits between 2003 and 2017.

Patient records were eligible if the patient was registered as HIV infected. Other inclusion criteria were to have started ART between 2003 and 2017, to be at least 18 years old at the start of treatment, to have at least one follow-up visit after the start of treatment. At inclusion, the following data were collected: sociodemographic data (date of birth, sex, nationality, education level, marital status), clinical data (HIV status, weight, height, WHO stage), standard biological data (AST, ALT, blood glucose, amylase, creatinine, blood cholesterol and triglyceride levels), virological data (plasma viral load, CD4 T-cell count). During follow-up, for the first two years, data were collected according to the following schedule: day 15, month 1 (M1), M3, M6, M9, M12, M15, M18, M21, and M24; then every 6 months until the 13th year after the start of treatment. For these different dates, the following data were collected: clinical data (therapeutic protocols, weight, WHO stage and information on therapeutic compliance), comorbidities (tuberculosis, viral hepatitis, malaria at each visit), vital status (death or loss to follow-up), virological data (viral load, CD4 T cell count from the 6th month of treatment and then every 6 months until the 14th year).

There were 18, 000 patients included at the first visit. The treatment regimens for the management of HIV patients in Congo are classified into two periods: Before (Guidelines published in 2014) and after (Guidelines published in 2018), as previously published. Regarding the first part of the Eval-co project on treatment failure, 6924 patients were included (reference n°13). We excluded patients under the age of 18 and those with no viral load data. For this second project, we excluded all patients who did not have a start date for antiretroviral therapy. At the end, 2,309 patients with the available data have been included for this study.

(Fig 1) shows the flowchart of the patients included in this study.

**Fig 1.**
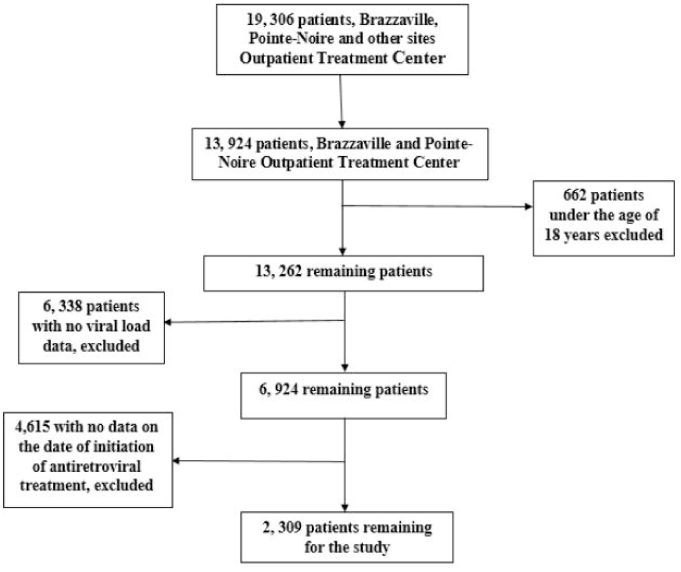
Flowchart of the patients for the study, eval-co historical cohort, 2003-2017, (n=2309).

### Presentation of study variables

The variables in our study included: socio-demographic data: sex (Male, Female); the patient’s age (in years), level of education (None, Primary, Secondary, University) and marital status (Single, Divorced, Married, Free union, Widowed). Clinical data: WHO clinical stage: stage I; II; III and IV [16]. Virological and immunological data: plasma viral load (in copies / ml) and CD4 T lymphocyte count (/ mm 3).

The variables on the survival analysis: the date of origin or initiation of the patient’s antiretroviral treatment, the point date set at December 31, 2017, the time of occurrence of the death event in years, the vital status of the patient or event of interest on the last news date or on the point date (0 = survival; 1 = death). In the event of death, we stopped the patient observation time on that date.

### Data analysis

We performed data analysis with Stata 15.0 software. The frequencies of socio-demographic characteristics (sex, age, marital status, level of education) were calculated. We used the Kaplan-Meier method to estimate the 14-year survival probabilities from the survival curve. The log-rank test was used to compare survival curves. The significance was set at 5% (p <0.05).

We used the Cox model to identify prognostic factors associated with patient death. In univariate analysis, each relevant explanatory variable identified in the review of the literature (sex, viral load, WHO clinical stage of the disease, CD4 number) was crossed with the explanatory variable “vital status” (1 = death; 0 = survival) [17–19]. The endpoint was death. The variables having a significant effect with a P value less than 0.05 were retained for the modeling. The significance level for integration into the final model was 0.05. We measured the strength of the association between the independent variable and the dependent variable by the hazard ratio, its 95% confidence interval, and the P-value.

### Ethical considerations

The research protocol has been approved by the ethics committee of the Faculty of Health Sciences (FSSA) of Marien NGOUABI University (UMNG) from Congo Brazzaville. We used routine data from all HIV AIDS monitoring and care centers in Congo. These data are transmitted to the CNLS-E (National Council for the Fight against HIV, AIDS and Epidemics) which centralizes all the country’s data through a national database. All data was anonymous. Only the team that participated in the research had access to the data. Permission to use the data was obtained from the Ministry of Health.

## Results

### Sociodemographic and clinical characteristics of patients

Table 2 shows that the median age of the patients was 49 years. Most of the patients were over 45 years old, i.e. 63.01%. Our study population was predominantly female, 68.56% versus 31.44% male. The majority of patients were single (26.89%) against 23.73% of free union couples. The most represented level of education was secondary level (58.86%). More than half of PLHIV had a CD4 count <200/mm^3^, i.e. 84.97%, and 80% had a viral load <1000 copies / ml. The majority of patients were in WHO clinical stage I, i.e. 69.34%.

**Table 2.** Sociodemographic and clinical characteristics of patients, Eval-co historical cohort 2003-2017, N = 2,309.

| Variable | Frenquency (N) | Percent (%) |
| --- | --- | --- |
| **Sex** |  |  |
| Male | 726 | 31.44 |
| Female | 1583 | 68.56 |
| **Age (in years)** |  |  |
| Mean ± SD | (49.19±11.09) | |
| Median [Q1, Q3] | 49[42-57] |  |
| Minimum; Maximum | 18;91 |  |
| 18-25 years | 73 | 3.16 |
| 26-35 years | 150 | 6.50 |
| 36-45 years | 631 | 27.33 |
| > 45 years | 1455 | 63.01 |
| **Marital Status** |  |  |
| Single | 621 | 26.89 |
| Divorced | 400 | 17.32 |
| Married | 362 | 15.68 |
| Free Union | 548 | 23.73 |
| Widower | 288 | 12.47 |
| Unspecified | 90 | 3.91 |
| **Level of education** |  |  |
| Unschoolled | 57 | 2.47 |
| Primary | 288 | 12.47 |
| Secondary | 1359 | 58.86 |
| University | 353 | 15.29 |
| Unspecified | 252 | 10.91 |
| **Number of CD4 T** |  |  |
| < 200 | 1962 | 84.97 |
| 200-350 | 109 | 4.72 |
| > 350 | 238 | 10.31 |
| **Viral Load (copies/ml)** |  |  |
| <1000 | 1847 | 80.00 |
| >1000 | 462 | 20.00 |
| **WHO Clinical Stage** |  |  |
| Stage I | 1601 | 69.34 |
| Stage II | 265 | 11.48 |
| Stage III | 276 | 11.95 |
| Stage IV | 167 | 7.23 |

### Overall survival and according to the different characteristics of the patients estimated by the kaplan-meier method, eval-co historical cohort 2003-2017, Overall survival curve

As shown in (Fig 2), under the baseline scenario, the overall survival at 14 years was 83.28% CI [78.29-87.22].

**Fig 2.**
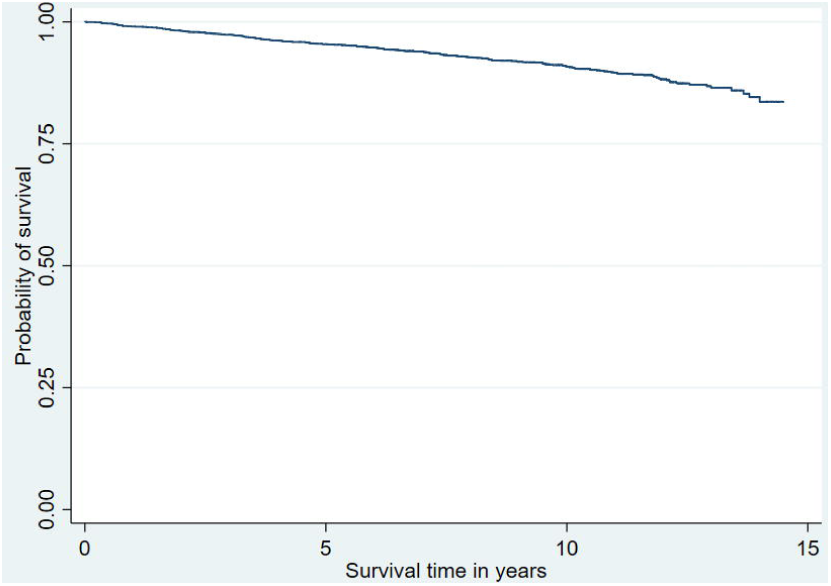
Kaplan Meier overall survival curve according to the baseline scenario.

### Worst case overall survival curve

As shown in (Fig 3), according to the worst-case scenario, if the lost to follow-up had died; the 14-year survival was 66.39% CI [62.14-70.29].

**Fig 3.**
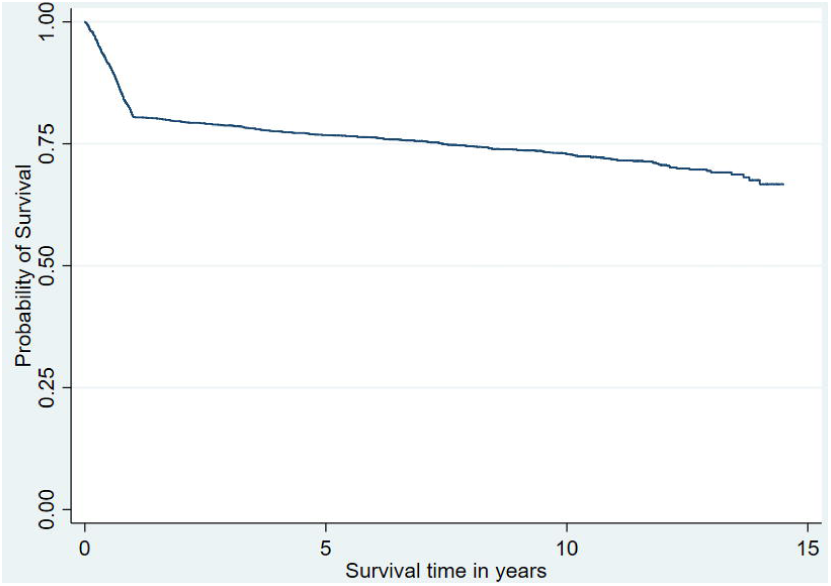
Kaplan Meier Worst Case Overall Survival Curve.

### Overall survival curve by sex

(Fig 4) shows us that: the comparison of survival curves after stratification of the sex variable shows that at 14 years old, men had a survival probability of 76.02% CI [63.31-84.84], lower than that of women, 86.98% CI [83.22-89.94]. This difference was significant, i.e. p <0.0001 (log-rank test).

**Fig 4.**
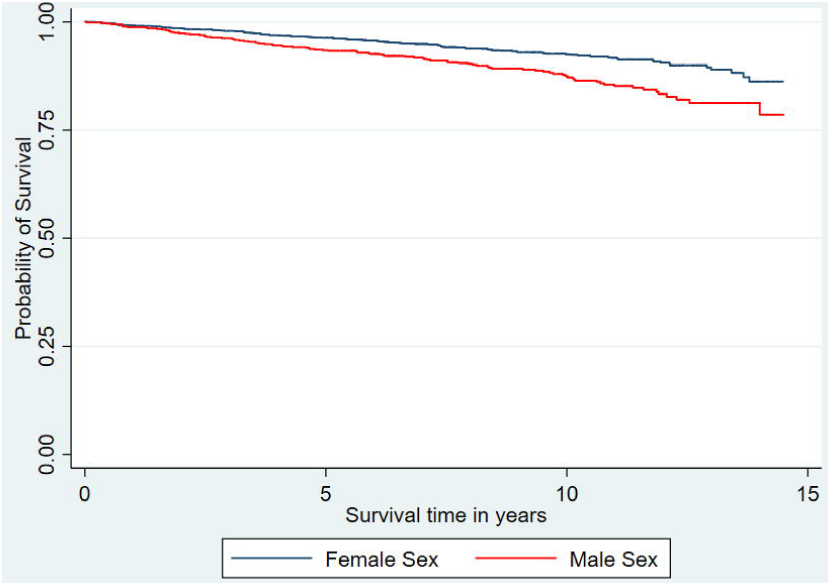
Kaplan Meier survival curve stratified by sex.

### Overall survival curve by CD4T

(Fig 5) shows us that: the comparison of survival curves after stratification of the variable CD4 T number shows that at 14 years, patients with a CD4 T number <200 / mm3 had a probability of survival of 82.03% [76.10-86.61. This is significantly lower than patients with a CD4 T count between 200-350 copies / ml and> 350 / mm3 with respectively: 79.12% [52.27-91.89]; 92.93% [87.21-96.15]. This difference was significant, i.e. p <0.02 (log-rank test).

**Fig 5.**
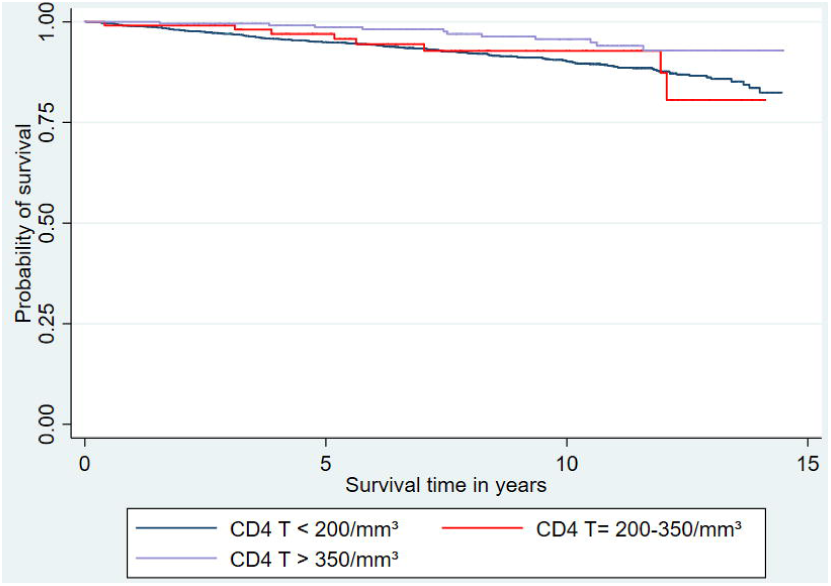
Kaplan Meier survival curve stratified by CD4 T.

### Overall survival curve by HIV viral load

(Fig 6) shows us that: the comparison of survival curves after stratification of the viral load variable shows that at 14 years, patients with a viral load> 1000 copies / ml had a probability of survival of 68.14% [53, 14-79, 23], significantly lower than patients with a viral load <1000 copies / ml, i.e. 87.74% [84.35-90.43]. This difference was significant, i.e. p <0.00001 (log-rank test).

**Fig 6.**
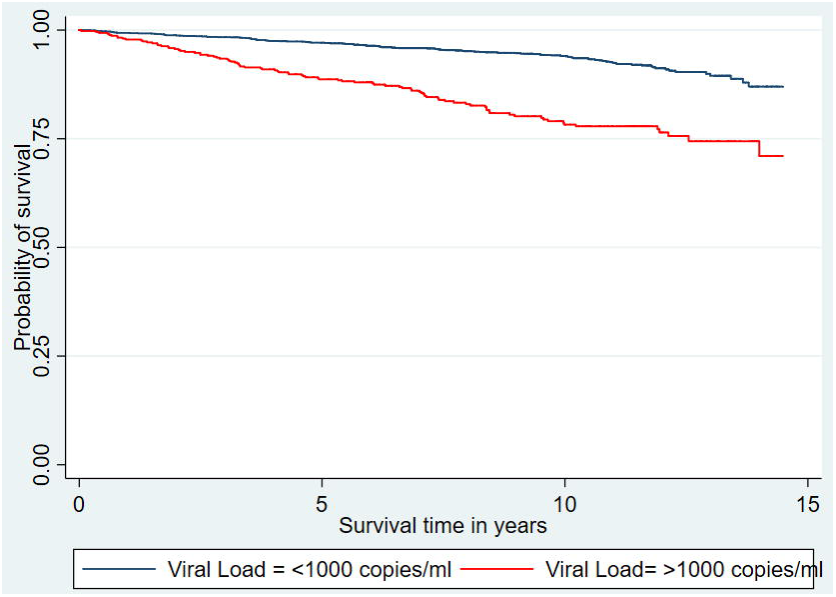
Kaplan Meier survival curve stratified by viral load.

### Overall survival curve by WHO clinical stage

(Fig 7) shows us: the comparison of survival curves after stratification of the WHO clinical stage variable shows that at 14 years, patients at WHO clinical stage = IV had a survival probability of 23.23% CI [14.05-33.76]. It was significantly lower than that of patients at clinical stage WHO = III, II and I with respectively: 34.83% [18.65-51.55]; 74.49% [42.13-90.45]; 93.82% [87.02-97.12]. This difference was significant, i.e. p <0.0001 (log-rank test).

**Fig 7.**
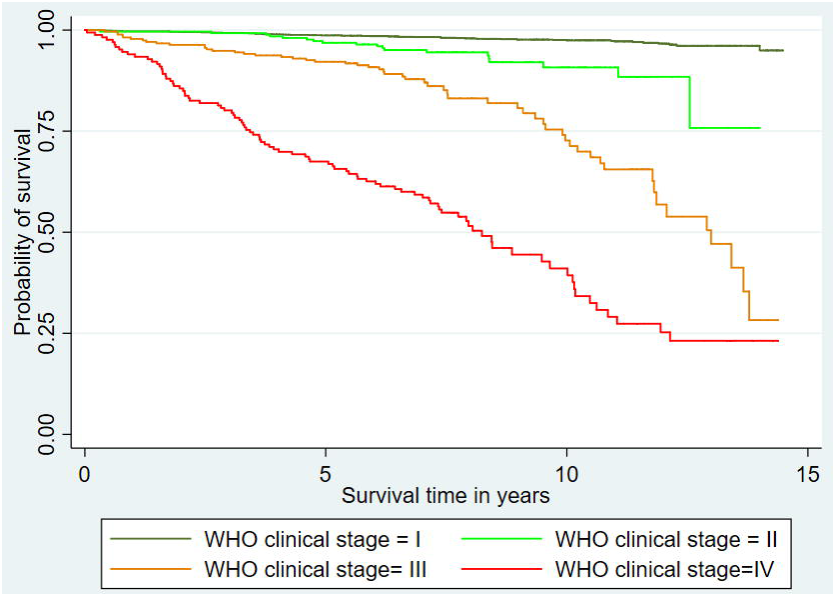
Overall Survival Curve by WHO Clinical Stage.

### Univariate and multivariate analysis stratified by cox regression of prognostic factors associated with death of PLHIV on art, historical eval-co cohort, 2003-2017, n = 2,309

Table 3 shows us that in univariate analysis, patient sex, WHO clinical stage of disease as well as viral load were strongly associated with the risk of death in HIV patients on ART. These variables also shown to be significantly associated with death in multivariate analysis. Cox’s regression analysis showed that men (AHR = 1.65, 95% CI [1.26-2.17]) had a significantly higher risk of death than women (P-value <0.0001).

**Table 3.** Multivariate analysis by cox regression of factors associated with death, Eval-co historical cohort 2003-2017, N = 2,309.

| Independent Variable | Univariate Analysis |  |  | Multivariate Analysis |  |  |
| --- | --- | --- | --- | --- | --- | --- |
|  | Unadjusted Hazard Ratio |  |  | Adjusted Hazard Ratio |  |  |
|  | HR | (CI 95%) | P-value | AHR | (CI 95%) | P-value |
| **Sex** |  |  |  |  |  |  |
| Female | 1 |  |  | 1 |  |  |
| Male | 1.74*** | [1.33-2.28] | 0.0001 | 1.65*** | [1.26-2.17] | 0.0001 |
| **WHO Clinical stage** |  |  |  |  |  |  |
| Stage I | 1 |  |  | 1 |  |  |
| Stage II | 3.85*** | [2.23-6.63] | 0.0001 | 4.07*** | [2.36-7.01] | 0.0001 |
| Stage III | 12.38*** | [8.30-18.49] | 0.0001 | 13.49*** | [8.99-20.27] | 0.0001 |
| Stage IV | 37.63*** | [26.11-54.24] | 0.0001 | 34.45*** | [23.74-50.0] | 0.0001 |
| **Number of CD4** |  |  |  |  |  |  |
| < 200/mm<sup>3</sup> | 1 |  |  | 1 |  |  |
| 200-350/mm<sup>3</sup> | 0.93 | [0.45-1.89] | 0.844 | 0.79 | [0.39-1.59] | 0.504 |
| >350/mm<sup>3</sup> | 0.44** | [0.24-0.81] | 0.008 | 0.91 | [0.49-1.68] | 0.754 |
| **Viral Load** |  |  |  |  |  |  |
| < 1000 copies/ml | 1 |  |  | 1 |  |  |
| > 1000 copies/ml | 3.14*** | [2.40-4.13] | 0.0001 | 2.56*** | [1.93-3.40] | 0.0001 |

CI: Confidence Interval; P-value < 0.05: \*; P-value < 0.01: \*\*; P-value < 0.001: \*\*\*.

HIV patients with advanced disease, especially stage II (AHR = 4.07, 95% CI [2.36-7.01]); stage III (AHR = 13.49, 95% CI [8.99-20.27]) and stage IV (AHR = 34.45, 95% CI [23.74-50]) were significantly associated with death compared to those in stage I of reference (p-value <0.0001). Patients with a viral load> 1000 copies / ml AHR = 2.56 95% CI [1.93-3.40]) were also significantly associated with death (p-value <0.0001).

## Discussion

Our work has shed light on the long-term survival of adult PLHIV on antiretroviral treatment between 2003-2017 in the Republic of Congo as well as the prognostic factors associated with death. In this retrospective cohort study, there were 212 deaths out of the 2,309 PLHIV included, for a death rate of 9.18%. Our descriptive results show a predominance of women compared to men (68.56% of women against 31.40% of men). This could be explain by the fact that women are more exposed to risky sexual practices given the financial precariousness that this layer faces. This is the case of sex workers where a single woman is been exposing to a high frequency of sexual relations with several men. They are at the same time the most at risk layer in the Republic of Congo [20].

The overall survival rate was 83% under the base case scenario and 66% under the worst scenario. Among the prognostic factors associated with death were: male sex advanced WHO clinical stage of the disease, and high viral load. The overall survival rate under the base case scenario was lower than that of the worst scenario by the simple fact that the lost to follow-up were considered to be dead. The fact that males were associated with the prognosis of death could be explained by several factors, the late diagnosis of the disease. This could lead to late management, an advanced clinical stage of the disease or a high viral load. Because in our study, the advanced clinical stage of the disease and the high viral load were also associated with the prognosis of death. Poor ARV adherence has also been a factor in treatment failure leading to a high rate of patient death in several African studies [21–25].

The probability of survival under the base scenario estimated to be 98% at one year, 97% at two years, 96% at three years, and 95% at 5 years. Our rates were slightly higher than those found in the DRC by LOANDO MBOYO A. et al. that were respectively 85% at one year, 82% at two years, 80% at three years and 78% at five years [26]. On the other hand, a study carried out in Cameroon by Kob Ye Sam et al. shows 91.3% survival at one year [27].

In the worst-case scenario, survival after two years estimated at 79%. In South Africa, Coetzee et al. measured survival on ARVs in adults at 86.3% at two years [28]. In Malawi, Zachariah et al. found 85% survival after two years [29].

In this study, according to the same scenario, the probability of survival at 5 years estimated at 76%. In Botswana, Bussmann H. and Collaborators, for the same duration found 79% (31). In Cameroon, Sieleunou and collaborators found 47% [30] against 75.4% in Senegal [31]. The male gender was a prognostic factor for mortality with a risk almost double that of the female sex. Our results corroborate those found in Nepal [32].

The final model on predictors of survival consisted of male gender, viral load, and WHO advanced clinical stage of the disease. [33]. Similar results have been found in several African studies [34–36]. The limits of the study: We did not include some possible predictors in the study due to variable insufficiency. These include, for example, hemoglobin level, therapeutic line, level of adherence to antiretroviral treatment, Tuberculosis coinfection, other opportunistic infections, body mass index, anemia, prophylactic treatment with cotrimoxazole.

## Conclusion

The long-term survival of PLHIV remains a concern according to the baseline scenario. These results could be much worse if all deaths were correctly reported. This survival would have been higher if the lost to follow-up were all taken for dead. In addition, being male, having an advanced clinical stage of the disease and a high viral load were the main prognostic factors significantly associated with death. A prospective study could consider modeling all of the prognostic factors identified in order to effectively fight HIV in the Republic of Congo.

In view of the results of this research, several actions are necessary by the institutions fighting against HIV / AIDS and the Ministry of Health in the Republic of Congo. Among which: Intensify mass awareness campaigns for HIV / AIDS screening. Ensure the availability of the necessary health-care inputs for the screening, monitoring and treatment of HIV AIDS. Organize an effective HIV AIDS data management system throughout the national territory for the completeness of information. Conduct closed and open cohort survival studies based on recent data.

## Data Availability

All relevant data are within the manuscript and its Supporting Information files

## Acknowledgments

We would like to thank all the staff of the National Council for the Fight against AIDS and Epidemics who facilitated the realization of our study. We would also like to thank all the researchers from Marien University who contributed to the proofreading of this research work.

## Author contributions

Project administration: GN. Conceptualization: GN AC MD AN. Analyzed the data: AN. Wrote the paper: GN AC AN MD.

## Supporting information

S1 dataset.survivalpatientsart.xls

S1 Fig 1. Flowchart of the patients for the study, eval-co historical cohort, 2003-2017, (n=2309).

S2 Fig 2. Kaplan Meier overall survival curve according to the baseline scenario.

S3 Fig 3. Kaplan Meier Worst Case Overall Survival Curve.

S4 Fig 4. Kaplan Meier survival curve stratified by sex.

S5 Fig 5. Kaplan Meier survival curve stratified by CD4 T.

S6 Fig 6. Kaplan Meier survival curve stratified by viral load

S7 Fig 7. Overall Survival Curve by WHO Clinical Stage

## References

1. HIV/AIDS HIV / AIDS [Internet]. [cited Sep 30 2021]. Available at: https://www.who.int/news-room/facts-in-pictures/detail/hiv-aids

2. Fact Sheet - Latest statistics on the state of the AIDS epidemic | UNAIDS [Internet]. [cited Sep 18 2021]. Available at: https://www.unaids.org/en/resources/fact-sheet.

3. HIV/AIDS n.d. https://www.who.int/news-room/fact-sheets/detail/hiv-aids (accessed September 30, 2021).

4. Barre-Sinoussi F. HIV / AIDS infection: the exemplary story of a resilient epidemic. MS Medicine Sci ISSN Pap 0767-0974 ISSN Number 1958-5381 Vol 34 N ° 6-7 P 499-500 [Internet]. 2018 [cited Sep 18 2021]; Available at: https://www.ipubli.inserm.fr/handle/10608/9804

5. Gruenais ME. A changing health system: the case of Cameroon. Autrepart-bondy PARIS-2004: 145–6.

6. Schneider H, Blaauw D, Gilson L, Chabikuli N, Goudge J. Health systems and access to antiretroviral drugs for HIV in Southern Africa: service delivery and human resources challenges. Reprod Health Matters. mai 2006; 14 (27): 12–23.

7. Cholera G-B, Cholera G-B. Joint United Nations Program on HIV / AIDS (UNAIDS) -WHO.

8. Gilks CF, Crowley S, Ekpini R, Gove S, Perriens J, Souteyrand Y, et al. The WHO public-health approach to antiretroviral treatment against HIV in resource-limited settings. Lancet Lond Engl. 5 août 2006; 368 (9534): 505–10.

9. Assessment and support of HAART in HIV-1 patients in Senegal - Google Scholar [Internet]. [cited Sep 18 2021]

10. C Seyler, X Anglaret, N Dakoury-Dogbo, E Messou, et al. Medium-term survival, morbidity and immunovirological evolution in HIV-infected adults receiving antiretroviral therapy, Abidjan, Côte d’Ivoire. Antivir Ther [Internet]. 1 oct 2003 [cited sept 18 2021]; 8 (5): 385–93. Available at: https://europepmc.org/article/med/14640385

11. Congo, Rep. - Survey of Seroprevalence and AIDS Indicators in Congo 2009 [Internet]. [cited Sep 30 2021]. Available at: https://microdata.worldbank.org/index.php/catalog/284812

12. WHO | HIV / AIDS in Congo n.d. https://www.who.int/countries/cog/areas/vih_sida/fr/ (accessed September 30, 2021).

13. Ndziessi G, Aloumba AJ, Essie DEM, Niama AC, Mbele FG, Diafouka M, et al. Factors Associated with Antiretroviral Treatments Failure among HIV-Positive Patients in Congo: A Retrospective Cohort Study. World J AIDS [Internet]. 25 dec 2020 [cited 18 sept 2021]; 10 (4): 201-14. Avalaible at: http://www.scirp.org/Journal/Paperabs.aspx?paperid=106130

14. Trop M. Therapeutic adherence of people living with HIV in 2009 at the outpatient treatment center in Brazzaville, Congo.Trop M. 2011; 71 (5): 487–91.

15. Elira Dokekias A, Atipo Galiba FO, Dzia Lepfoundzou Bokilo A, Ntsimba P, Nsitou MB. Evaluation of antiretroviral therapy in adults infected with HIV, followed in the hematology service of the CHU of Brazzaville, Congo. Bull Company Pathol Exot. 2008; 101 (2): 109–112.

16. Ghoma Linguissi LS, Lucaccioni V, Bates M, Zumla A, Ntoumi F. Achieving sustainable development goals for HIV/AIDS in the Republic of the Congo — Progress, obstacles and challenges in HIV/AIDS health services. Int J Infect Dis [Internet]. 1 dec 2018 [cited 18 sept 2021]; 77: 107–12. Available at: https://www.sciencedirect.com/science/article/pii/S1201971218345557

17. Medicalcul - Stades cliniques OMS du SIDA ∼ Infectiologie [Internet]. [cited 3 oct 2021]. Available at: http://medicalcul.free.fr/omssida.html

18. Zhang G, Gong Y, Wang Q, Deng L, Zhang S, Liao Q, et al. Outcomes and factors associated with survival of patients with HIV/AIDS initiating antiretroviral treatment in Liangshan Prefecture, southwest of China: A retrospective cohort study from 2005 to 2013. Medicine (Baltimore) [Internet]. juill 2016 [cited 20 nov 2021]; 95 (27). Available at: https://www.ncbi.nlm.nih.gov/labs/pmc/articles/PMC5058800/

19. Nacarapa E, Verdu ME, Nacarapa J, Macuacua A, Chongo B, Osorio D, et al. Predictors of attrition among adults in a rural HIV clinic in southern Mozambique: 18-year retrospective study. Sci Rep [Internet]. sept 9 2021 [cited 20 nov 2021]; 11 (1): 17897. Available at: https://www.nature.com/articles/s41598-021-97466-2

20. Trickey A, May MT, Vehreschild J-J, Obel N, Gill MJ, Crane HM, et al. Survival of HIV-positive patients starting antiretroviral therapy between 1996 and 2013: a collaborative analysis of cohort studies. Lancet HIV [Internet]. 1 août 2017 [cited 20 nov 2021]; 4 (8): e349–56. Available at: https://www.thelancet.com/journals/lanhiv/article/PIIS2352-3018(17)30066-8/fulltext

21. (CNLS) of the Republic of Congo. Behavioral survey coupled with hiv serology among sex workers, men who have sex with men and detainees in the republic of the congo final report [Internet]. BRAZZAVILLE; 2012. Available on: http://www.plateformeelsa.org/wpcontent/uploads/2017/02/Rapport_PS_MSM_Detenus_Congo.pdf

22. Study of factors linked to adherence to antiretroviral therapy in patients followed at the HIV / AIDS Management Unit of Dschang District Hospital, Cameroon [Internet]. [cited 3 Oct 2021]. Available at: https://panafrican-med-journal.com/content/article/12/55/full/

23. Ollivier F, N’Kam M, Midoungue C, Rey J-L. Study on adherence to antiretroviral treatment at the Yaoundé University Hospital Center (Cameroon). Public Health (Bucur) [Internet].2005 [cited Oct 3 2021]; Vol. 17 (4): 559–68. Available on: https://www.cairn.info/revue-sante-publique-2005-4-page-559.htm

24. Study on adherence to antiretroviral treatment at the Yaounde University Hospital Center (Cameroon) | Cairn.info [Internet]. [cited Oct 3 2021]. Available at: https://www.cairn.info/revue-sante-publique-2005-4-page-559.htm

25. Adherence to antiretroviral treatment: African particularities - EM consults [Internet]. [cited Oct 3 2021]. Available at: https://www.em-consulte.com/article/54032

26. Observance to antiretroviral treatment in the rural region of the Democratic Republic of Congo: a cognitive dissonance [Internet]. [cited Oct 3 2021]. Available on: https://www.ncbi.nlm.nih.gov/pmc/articles/PMC6488246/

27. Mboyo AL. Effectiveness after 5 years of an access to antiretroviral therapy project in the DRC.

28. Anouar D, Ye K, Iii S, Billong S, Fokam J, Momo P, et al. Survival of patients on antiretroviral therapy in Cameroon. 7 mai 2019.

29. Coetzee D, Hildebrand K, Boulle A, Maartens G, Louis F, Labatala V, et al. Outcomes after two years of providing antiretroviral treatment in Khayelitsha, South Africa. Aids. 2004; 18 (6): 887–95.

30. Zachariah R, Fitzgerald M, Massaquoi M, Pasulani O, Arnould L, Makombe S, et al. Risk factors for high early mortality in patients on antiretroviral treatment in a rural district of Malawi. Aids. 2006; 20 (18): 2355–60.

31. Bussmann H, Wester CW, Ndwapi N, Grundmann N, Gaolathe T, Puvimanasinghe J, et al. Five Year Outcomes of Initial Patients Treated in Botswana’s National Antiretroviral Treatment Program. AIDS Lond Engl [Internet]. nov 12 2008 [cited 18 sept 2021]; 22 (17): 2303–11. Available at: https://www.ncbi.nlm.nih.gov/pmc/articles/PMC2853026/

32. Sieleunou I, Souleymanou M, Schönenberger A-M, Menten J, Boelaert M. Determinants of survival in AIDS patients on antiretroviral therapy in a rural centre in the Far-North Province, Cameroon. Trop Med Int Health. 2009; 14 (1): 36–43.

33. Mortality and causes of death in adults receiving highly act…_J: AIDS [Internet]. [cited 18 sept 2021]. Available at: https://journals.lww.com/aidsonline/Fulltext/2006/05120/Mortality_and_causes_of_death_in_adults_receiving.12.aspx

34. Bhatta L, Klouman E, Deuba K, Shrestha R, Karki DK, Ekstrom AM, et al. Survival on antiretroviral treatment among adult HIV-infected patients in Nepal: a retrospective cohort study in far-western Region, 2006–2011. BMC Infect Dis. 2013; 13 (1):

35. Predictors of mortality in HIV-infected patients starting antiretroviral therapy in a rural hospital in Tanzania | BMC Infectious Diseases | Full Text [Internet]. [cited 3 oct 2021]. Available at: https://bmcinfectdis.biomedcentral.com/articles/10.1186/1471-2334-8-52

36. Biset Ayalew M. Mortality and its predictors among HIV infected patients taking antiretroviral treatment in Ethiopia: a systematic review. AIDS Res Treat. 2017.

37. Biadgilign S, Reda AA, Digaffe T. Predictors of mortality among HIV infected patients taking antiretroviral treatment in Ethiopia: a retrospective cohort study. AIDS Res Ther. 2012; 9 (1): 1–7.

38. Joseph N, Sinha U, Tiwari N, Ghosh P, Sindhu P. Prognostic factors of mortality among adult patients on antiretroviral therapy in India: A hospital based retrospective cohort study. BioMed Res Int. 2019; 2019.

